# Dystonia-Parkinsonism Gene Variants in Individuals with Parkinsonism and Brain Scans without Evidence for Dopaminergic Deficit (SWEDD)

**DOI:** 10.1101/2022.03.21.22271781

**Authors:** Kelly N. H. Nudelman, Yajing Xiong, Lola Cook, Jeanine Schulze, Marco Abreu, Kenneth Marek, Andrew Singleton, Tatiana Foroud, Lana M. Chahine, The Parkinson’s Progression Markers Initiative

## Abstract

**Objective:** To investigate the genetic etiology of dystonia-parkinsonism among individuals with parkinsonism who have dopamine transporter (DAT) SPECT scans without evidence of dopaminergic deficits (SWEDD).

**Methods:** Data for this case-control study were from the Parkinson’s Progression Markers Initiative (PPMI) cohort, a multisite observational study. The sample analyzed included participants with whole genome sequencing (WGS) who were diagnosed as Parkinson’s disease (PD, n=421), SWEDD (n=64) or healthy controls (HC, n=196). WGS data were analyzed to identify rare pathogenic variants in 30 dystonia-parkinsonism genes. Rare variants were prioritized for those present in individuals with SWEDD, but not in HC, reported pathogenicity in the literature and genetic databases, and with data from programs analyzing variant conservation and effect on protein function. For each SWEDD case with predicted or reported pathogenic variant(s), demographic, clinical, and imaging data were reviewed.

**Results:** Eight of the 64 SWEDD participants (12.5%) had a pathogenic variant in one or more dystonia-parkinsonism genes, including *PRKRA, SGCE, GLB1, ADCY5, SLC6A3*, and *GCDH*. Notably, one case had a heterozygous variant in *SGCE*, p.R263H, which is predicted pathogenic but currently classified as a variant of uncertain significance due to insufficient evidence.

**Conclusions:** Pathogenic variants in parkinsonism-dystonia genes occurred in more than 10% of SWEDD cases in the PPMI cohort, supporting the importance of considering dystonia disorders in the differential diagnosis of PD, particularly in the case of participants with normal DAT SPECT scans. Analysis of genetic variants in parkinsonism-dystonia genes in SWEDD cases can provide information which will improve clinical diagnosis and management.

**KEY MESSAGES:** *What is already known on this topic:* Individuals with parkinsonism with scans without evidence of dopamine deficiency (SWEDD) constitute up to 16% of subjects with early parkinsonism referred for inclusion in multicenter observational studies and randomized clinical trials; however, the majority of these individuals do not have Parkinson’s disease. Dystonic disorders are high on the list of differential diagnoses for individuals with parkinsonism and SWEDD, and an increasing number of studies support genetic etiologies for dystonic disorders. What proportion of individuals with parkinsonism and SWEDD referred for research studies might carry pathogenic variants in dystonia-parkinsonism related genes is not known.

*What this study adds:* This study identified predicted or reported pathogenic variants in dystonia-parkinsonism related genes in more than 10% of SWEDD cases in the PPMI cohort, supporting the utility of evaluating dystonia-parkinsonism related genetic etiology in SWEDD cases.

*How this study might affect research, practice, or policy:* This study supports the importance of genetic analyses in SWEDD cases which could improve clinical diagnosis and patient management.

## INTRODUCTION

Parkinsonism, a common movement disorder in older adults, is commonly caused by idiopathic Parkinson’s disease (PD). However, many individuals with parkinsonism are misdiagnosed as having PD.^1^ Dopamine transporter (DAT) single-photon emission computerized tomography (SPECT) imaging improves diagnostic accuracy in parkinsonism; reduced DAT binding indicates nigrostriatal denervation as seen in PD and other parkinsonian disorders. A normal DAT SPECT essentially rules out a PD diagnosis.^2^ Up to 16% of individuals with early parkinsonism referred for inclusion in multicenter observational studies and randomized clinical trials (RCTs) have scans without evidence of dopamine deficiency (SWEDD).^3-5^

Among individuals with SWEDD in the research setting, <5% later have an abnormal DAT SPECT and receive a diagnosis of PD or another parkinsonian disorder.^3^ In the remaining individuals, scans continue to be normal; it is unlikely that these individuals have a neurodegenerative parkinsonian disorder. Dystonic disorders are frequently a differential diagnosis,^3^ as many patients have dystonia on exam, and dystonic tremor may mimic PD rest tremor. Genetic causes for dystonia are increasingly recognized.^6^ A genetic diagnosis in an individual with dystonia could alter prognostic counseling and patient management.^7^

In light of the reported contribution of dystonic disorders to the clinical presentation of individuals with parkinsonism and SWEDD, we examined the prevalence of rare, predicted or reported pathogenic variants in dystonia-parkinsonism related genes among a research cohort of individuals with parkinsonism and normal DAT imaging. We hypothesized that we would identify pathogenic variants in dystonia-parkinsonism related genes in SWEDD cases, which would provide insight into participant diagnosis.

## METHODS

This was a case-control study comparing the prevalence of pathogenic variants in dystonia-parkinsonism genes in individuals with SWEDD compared to HC and PD.

Data were from the Parkinson Progression Markers Initiative (PPMI), an international multisite prospective observational study investigating biomarkers for PD. Study methodology can be found at ppmi-info.org. Data were obtained from the Laboratory of NeuroImaging (LONI, www.loni.usc.edu) and are available for download by qualified investigators.

### Patient Consent and Study Approval

Written informed consent was obtained from all PPMI participants for consent to research, including genetic research. The PPMI study was conducted in accordance with the ethical standards of the Helsinki Declaration of 1975. Secondary analyses for this project were approved under the Indiana University IRB protocol 12743.

### Sample

The sample included individuals enrolled in PPMI with PD diagnosed within the prior two years and untreated for PD at baseline. An abnormal DAT SPECT scan (defined by visual read) was required for inclusion in the PD cohort; individuals with normal DAT SPECT were eligible to be enrolled in the SWEDD cohort. Healthy controls (HC) with normal DAT SPECT were also enrolled.

Of the 570 screened participants with parkinsonism, 423 with abnormal DAT imaging were enrolled in the PD cohort. 81 had SWEDD, of which 64 enrolled. Final analysis sample included 64 SWEDD cases, 421 PD cases, and 196 HC with whole genome sequencing (WGS); participants were analyzed according to their enrollment cohort (n=681; see Figure 1).

**Figure 1.**
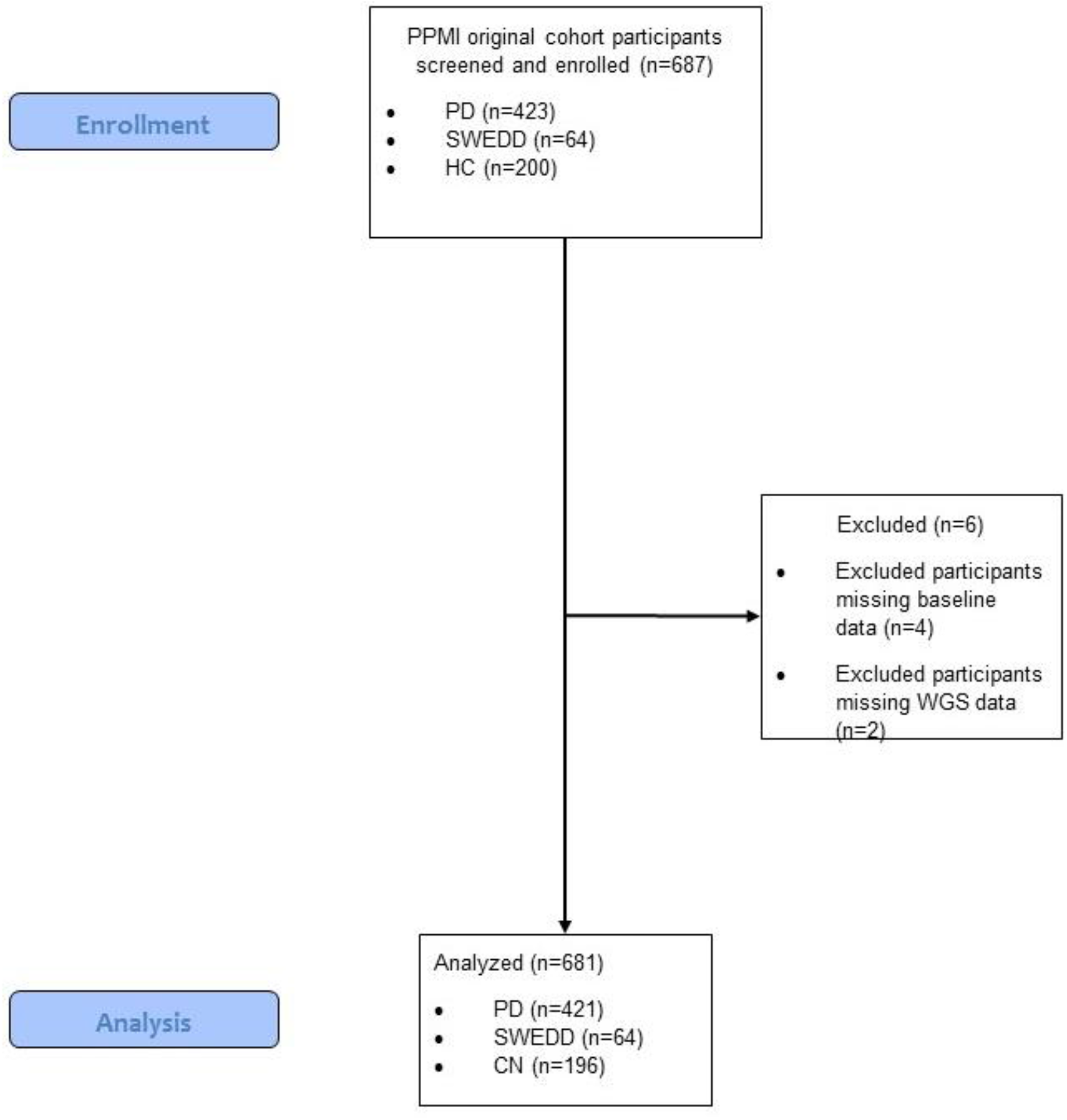
Consort Diagram. The PPMI original cohort (Parkinson’s disease (PD), SWEDD, and healthy controls (HC)) included 687 subjects screened and enrolled; the sample included 423 PD, 64 SWEDD, and 200 HC. Of these, four HC were excluded for missing baseline data, and another two PD were excluded for missing whole genome sequencing (WGS) data, yielding a final analysis sample of n=681.

### Genetic Analyses

WGS variant-level data for 30 dystonia-parkinsonism genes were reviewed for occurrence in SWEDD cases but not HC, minor allele population frequency and reported and predicted pathogenicity. (For details see Supplemental Methods).

### Case Reviews

For each case carrying predicted or reported pathogenic variant(s), a review of clinical and imaging data collected at the screening/baseline and one- and two-year follow-up assessments was conducted to construct the case histories based on all available data, and to account for the expected phenotype for pathogenic variants in each gene (for more details on clinical and imaging methods see Supplemental Methods). Sex and age at first visit are presented in aggregate, and age at onset presented in ranges, to reduce potentially identifying information for participants.

### Statistical Analysis

Demographic variables age at study baseline, sex, years of education, race, and ethnicity were assessed for differences between SWEDD, PD, and HC groups. Analyses were performed in SPSS version 27 (SPSS Statistics 27, IBM Corporation, Somers, NY) using ANOVA for continuous variables or Pearson Chi-Square tests for nominal variables.

## RESULTS

Demographics are summarized in Table 1; no significant differences were identified across diagnostic groups.

**Table 1.**
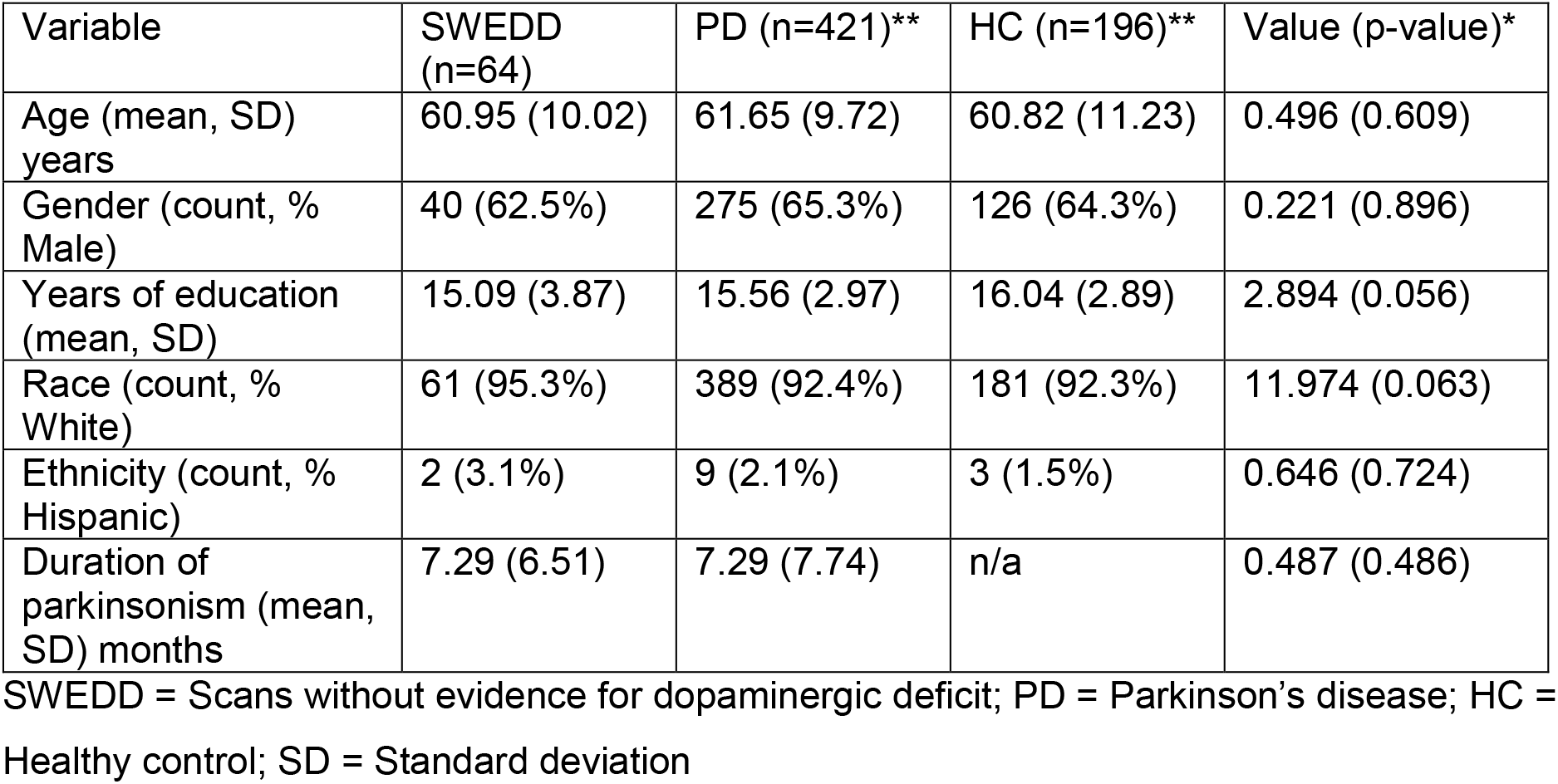
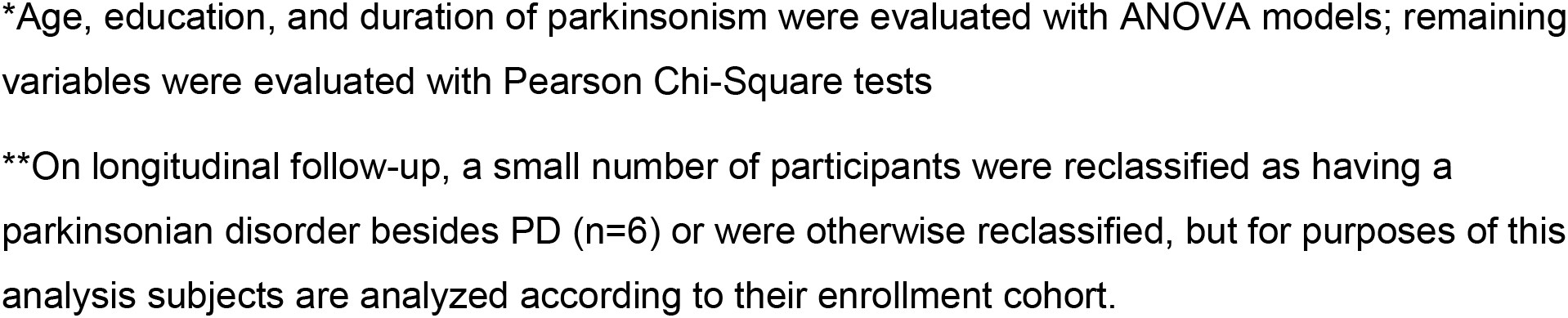
Sample Demographics

| Variable | SWEDD (n=64) | PD (n=421)** | HC (n=196)** | Value (p-value)* |
| --- | --- | --- | --- | --- |
| Age (mean, SD) years | 60.95 (10.02) | 61.65 (9.72) | 60.82 (11.23) | 0.496 (0.609) |
| Gender (count, % Male) | 40 (62.5%) | 275 (65.3%) | 126 (64.3%) | 0.221 (0.896) |
| Years of education (mean, SD) | 15.09 (3.87) | 15.56 (2.97) | 16.04 (2.89) | 2.894 (0.056) |
| Race (count, % White) | 61 (95.3%) | 389 (92.4%) | 181 (92.3%) | 11.974 (0.063) |
| Ethnicity (count, % Hispanic) | 2 (3.1%) | 9 (2.1%) | 3 (1.5%) | 0.646 (0.724) |
| Duration of parkinsonism (mean, SD) months | 7.29 (6.51) | 7.29 (7.74) | n/a | 0.487 (0.486) |

SWEDD = Scans without evidence for dopaminergic deficit; PD = Parkinson's disease; HC = Healthy control; SD = Standard deviation
\*Age, education, and duration of parkinsonism were evaluated with ANOVA models; remaining variables were evaluated with Pearson Chi-Square tests
\*\*On longitudinal follow-up, a small number of participants were reclassified as having a parkinsonian disorder besides PD (n=6) or were otherwise reclassified, but for purposes of this analysis subjects are analyzed according to their enrollment cohort.

We identified a final list of eight putative or reported pathogenic variants in six genes: protein activator of interferon induced protein kinase EIF2AK2 (*PRKRA)*, solute carrier family 6 member 3 (*SLC6A3)*, galactosidase beta 1 *(GLB1)*, glutaryl-CoA dehydrogenase *(GCDH)*, adenylate cyclase 5 *(ADCY5)*, and sarcoglycan epsilon (*SGCE)*. These variants were not found in any HC participants. Variants are listed and annotated in Table 2.

**Table 2.**
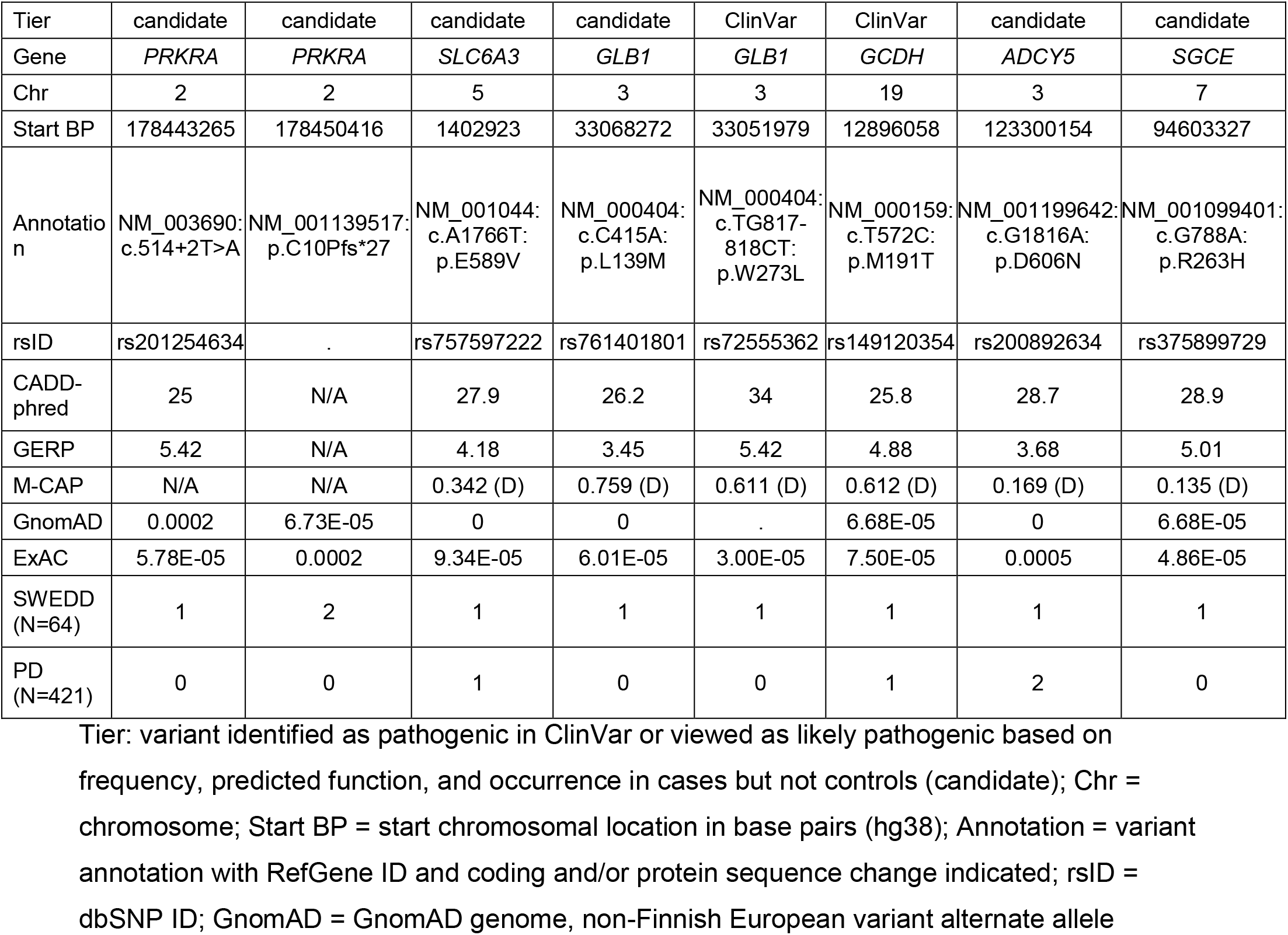
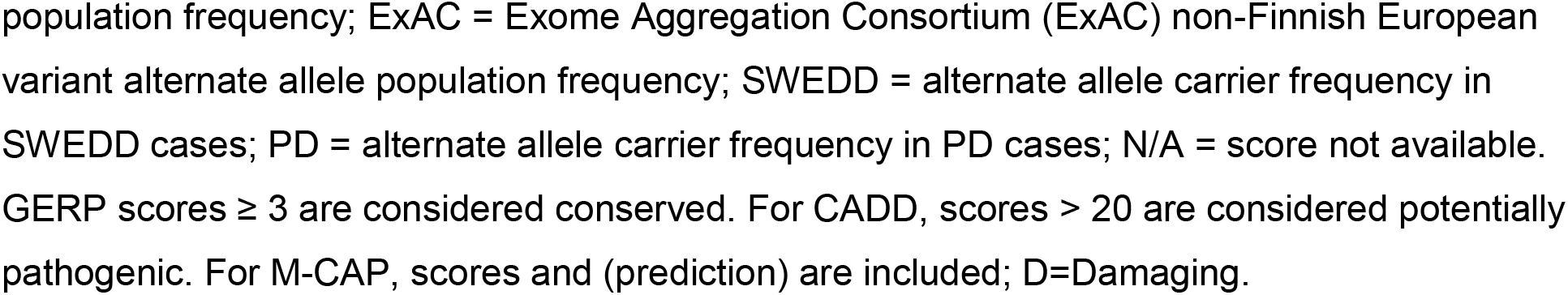
SWEDD Variants in Dystonia-Parkinsonism Genes

| Tier | candidate | candidate | candidate | candidate | ClinVar | ClinVar | candidate | candidate |
| --- | --- | --- | --- | --- | --- | --- | --- | --- |
| Gene | *PRKRA* | *PRKRA* | *SLC6A3* | *GLB1* | *GLB1* | *GCDH* | *ADCY5* | *SGCE* |
| Chr | 2 | 2 | 5 | 3 | 3 | 19 | 3 | 7 |
| Start BP | 178443265 | 178450416 | 1402923 | 33068272 | 33051979 | 12896058 | 123300154 | 94603327 |
| Annotation | NM_003690:<br>c.514+2T>A | NM_001139517:<br>p.C10Pfs*27 | NM_001044:<br>c.A1766T:<br>p.E589V | NM_000404:<br>c.C415A:<br>p.L139M | NM_000404:<br>c.TG817-818CT:<br>p.W273L | NM_000159:<br>c.T572C:<br>p.M191T | NM_001199642:<br>c.G1816A:<br>p.D606N | NM_001099401:<br>c.G788A:<br>p.R263H |
| rsID | rs201254634 | . | rs757597222 | rs761401801 | rs72555362 | rs149120354 | rs200892634 | rs375899729 |
| CADD-phred | 25 | N/A | 27.9 | 26.2 | 34 | 25.8 | 28.7 | 28.9 |
| GERP | 5.42 | N/A | 4.18 | 3.45 | 5.42 | 4.88 | 3.68 | 5.01 |
| M-CAP | N/A | N/A | 0.342 (D) | 0.759 (D) | 0.611 (D) | 0.612 (D) | 0.169 (D) | 0.135 (D) |
| GnomAD | 0.0002 | 6.73E-05 | 0 | 0 | . | 6.68E-05 | 0 | 6.68E-05 |
| ExAC | 5.78E-05 | 0.0002 | 9.34E-05 | 6.01E-05 | 3.00E-05 | 7.50E-05 | 0.0005 | 4.86E-05 |
| SWEDD (N=64) | 1 | 2 | 1 | 1 | 1 | 1 | 1 | 1 |
| PD (N=421) | 0 | 0 | 1 | 0 | 0 | 1 | 2 | 0 |

Tier: variant identified as pathogenic in ClinVar or viewed as likely pathogenic based on frequency, predicted function, and occurrence in cases but not controls (candidate); Chr = chromosome; Start BP = start chromosomal location in base pairs (hg38); Annotation = variant annotation with RefGene ID and coding and/or protein sequence change indicated; rsID = dbSNP ID; GnomAD = GnomAD genome, non-Finnish European variant alternate allele
population frequency; ExAC = Exome Aggregation Consortium (ExAC) non-Finnish European variant alternate allele population frequency; SWEDD = alternate allele carrier frequency in SWEDD cases; PD = alternate allele carrier frequency in PD cases; N/A = score not available. GERP scores $\geq 3$ are considered conserved. For CADD, scores $> 20$ are considered potentially pathogenic. For M-CAP, scores and (prediction) are included; D=Damaging.

Results of the patient- and clinician-reported outcomes administered as part of participants’ study assessments are shown in Table 3. Six patients were female and two male; age at first study visit ranged from 49 to 68 years. A description of the eight SWEDD cases with putative or reported pathogenic variant(s) follows.

**Table 3.** Case Demographics and Clinical Information

| Case# | Gene(s) | Variant type(s) | Race | Lowest Putamen SBR-Adj | MDS-UPDRS total score | MDS-UPDRS part 1 total score | MDS-UPDRS part 2 total score | MDS-UPDRS part 3 total score | UPSIT | MOCA | STAI total score | SCOPA - AUT | GDS | ESS | RBDSQ |
| --- | --- | --- | --- | --- | --- | --- | --- | --- | --- | --- | --- | --- | --- | --- | --- |
| 1 | PRKRA | Splicing | White | 0.544 | 37 | 12 | 9 | 16 | 25 | 25 | 99 | 10 | 9 | 16 | 5 |
| 2 | PRKRA | Frameshift insertion | White | 1.209 | 17 | 12 | 3 | 2 | 38 | 27 | 82 | 23 | 6 | 14 | 6 |
| 3 | SLC6A3, PRKRA | NS-SNV, frameshift insertion | White | 0.801 | 29 | 9 | 2 | 18 | 35 | 29 | 59 | 7 | 0 | 7 | 4 |
| 4 | GLB1 | NS-SNV | White | 0.904 | 26 | 8 | 3 | 15 | 36 | 29 | 65 | 9 | 2 | 8 | 5 |
| 5 | GLB1 | NS-SNV | White | 0.716 | 45 | 14 | 12 | 19 | 25 | 27 | 60 | 19 | 1 | 9 | 6 |
| 6 | GCDH | NS-SNV | White | 0.930 | 14 | 2 | 7 | 5 | 35 | 30 | 40 | 6 | 0 | 14 | 2 |
| 7 | ADCY5 | NS-SNV | White | 0.728 | 8 | 1 | 2 | 5 | 35 | 27 | 48 | 5 | 0 | 2 | 2 |
| 8 | SGCE | NS-SNV | White | 0.819 | 37 | 11 | 14 | 12 | 36 | 28 | 69 | 16 | 2 | 11 | 1 |

Lowest putamen SBR-Adj = lowest putamen signal-to-noise-ratio from SPECT DAT scan divided by expected value for age and sex as in Chahine et al., 2021;<sup>8</sup> MDS-UPDRS = Movement Disorders Society Unified Parkinson's Disease Rating Scale; UPSIT = University of Pennsylvania Smell Identification Test; MoCA = Montreal Cognitive Assessment; STAI = State-Trait Anxiety Inventory; SCOPA-AUT = Scales for Outcomes in PARKinson's disease-Autonomic; GDS = Geriatric Depression Scale; ESS = Epworth Sleepiness Scale; RBDSQ= REM sleep behavior disorder questionnaire; NS-SNV = nonsynonymous single nucleotide variant.

**Case 1**. This participant was heterozygous for a rare *PRKRA* splice donor variant (NM_003690 c.514+2T>A) near exon 5. While this variant does not currently meet American College of Medical Genetics (ACMG) criteria for pathogenicity, this variant is within 2 base pairs of a splice site in a gene where loss of function is a known disease mechanism, and computational predictions support pathogenicity.

Homozygous variants in the *PRKRA* gene have been associated with DYT-PRKRA (DYT16). The *PRKRA* gene encodes a protein kinase that mediates the effects of interferon response to viral infections and has been implicated in signal transduction, cell differentiation and proliferation, and apoptotic pathways, though the exact causal mechanism of dystonia symptoms is unclear.^9^ The most common reported *PRKRA* pathogenic variant is p.P222L, which segregates in an autosomal recessive (AR) inheritance pattern with the disorder in Brazilian patients.^9^ Several heterozygous variants have been identified in the *PRKRA* gene in a population of adult-onset focal or segmental dystonia;^10^ however, it is unclear if these variants are contributing to disease in these patients. This splicing variant, c.514+2T>A, has not previously been reported in the literature for dystonia, though it is present in dbSNP (rs201254634).

DYT-PRKRA (DYT16) is characterized by early-onset dystonia parkinsonism, including gait abnormalities, dysphagia, dystonia, torticollis, and opisthotonic posturing. Some patients also display bradykinesia, tremor and rigidity, delayed development, and speech and language impairment ^9^.

This case was right-handed with 12 years education and parkinsonism onset between 60-65 years. This patient had medical history of coronary artery disease & myocardial infarction s/p stent and obstructive sleep apnea (OSA), with no family history of PD. Presenting symptoms included resting tremor and rigidity with the right hand affected more than the left. On the initial examination, bradykinesia was also noted. This participant had mild cognitive impairment with a MOCA of 25. Of note, olfactory testing revealed mild hyposmia. Case 1 had later symptom onset than previously reported for DYT-PRKRA (DYT16), but case presentation was otherwise consistent with the described phenotype.

**Case 2**. This case was heterozygous for a rare frameshift variant introducing a premature stop codon in the first exon of the *PRKRA* gene at amino acid 27 (NM_001139517 c.27_28insCCCTTCTCGCCCTGTCCCAGAGCAGGCACCGCCGAGGCCCCGCCGCTGGAGC GCGAGGACAG, p.C10Pfs*27). This variant is likely to be deleterious; however, as stated for case 1, the effect of heterozygous functional variants in *PRKRA* is unclear. Interestingly, Seibler and colleagues reported a juvenile onset dystonia patient heterozygous for a different novel frameshift variant in *PRKRA* that also introduced a premature stop codon (p.H89fsX20),^11^ supporting the possibility of dosage effects for *PRKRA* heterozygous pathogenic variants in dystonia.

This participant was right-handed with 16 years education and parkinsonism onset between 45-50 years. Patient medical history included diverticulitis and large bowel resection. There was no family history of PD. The exam showed bilateral upper extremity rigidity and bradykinesia; otherwise, the neurological exam was unremarkable. The participant had a SCOPA-AUT score of 23 with evidence of urological and gastrointestinal autonomic dysfunction. This patient’s motor symptoms are not inconsistent with DYT-PRKRA (DYT16), though with much later onset compared to patients homozygous for a pathogenic *PRKRA* variant.

**Case 3**. This participant was heterozygous for a rare coding variant in exon 3 of the *SLC6A3* gene (NM_001044 c.A1766T, p.E589V). This variant is within 2 base pairs of a splice site in a gene where loss of function is a known mechanism of disease (ACMG PVS1 very strong criteria), the variant is present at extremely low frequency in GnomAD population databases (PM2 moderate criteria), and computational predictions support pathogenic function (PP3 supporting criteria); therefore, this variant meets the ACMG criteria for pathogenicity. Interestingly, this participant was also heterozygous for a rare frameshift insertion in the *PRKRA* gene, p.C10Pfs*27, also present in Case 2, though these two cases are not related.

Homozygous or compound heterozygote pathogenic variants in *SLC6A3* gene cause infantile parkinsonism-dystonia-1 (PKDYS1); however, it is unclear whether heterozygous *SLC6A3* variants are disease-causing. It is also currently unclear whether heterozygous variants in *PRKRA* are disease-causing, though there is some evidence in the literature to suggest that heterozygous functional variants do occur in other cohorts of dystonia patients.^10^ Symptoms of both diseases typically include bradykinesia and tremor.

This case was left-handed with 21 years education and parkinsonism onset between 50-55 years. Main symptoms were tremors and bradykinesia, though with a much later age of onset than reported for PKDYS1 or DYT-PRKRA (DYT16). Preexisting medical conditions included hypothyroidism, depression, and arthritis, with no family history of PD.

**Case 4**. This participant was heterozygous for a rare coding variant in exon 4 of the *GLB1* gene (NM_000404 c.C415A, p.L139M), which is predicted to be deleterious. This variant is likely pathogenic according to ACMG criteria – it occurs in an exonic hotspot (PM1 criteria) and is at very low frequency in population databases (PM2). This is a novel missense variant at a position where another variant has been classified likely pathogenic (PM5), occurs in a gene that has a low rate of benign missense variation and in which missense variants are a common mechanism of disease (PP2), and computational predictions support deleterious variant function (PP3). However, *GLB1* shows AR inheritance for GM1-gangliosidosis and muclpolysaccharidosis type IVB, so it is unclear if this likely pathogenic variant contributes to disease presentation in this case.

Homozygous mutations in the *GLB1* gene cause GM1 gangliosidosis, a lysosomal storage disease with symptoms including dystonia, akinetic-rigid parkinsonism, and multiple bone issues. MRI shows bilateral putamen T2 hyperintensities, which in patients with this disease suggest accumulation of GM1 ganglioside to toxic levels. Patients with GM1 gangliosidosis can also present with abnormal morphology of their long bones.

This case was right-handed with 12 years education and parkinsonism onset between 45-50 years. This case had a medical history of obstructive sleep apnea, gastrointestinal reflux disease, and depression, but reported no orthopedic surgeries; this case had a first-degree relative with diagnosed PD. The examination showed parkinsonism with no other abnormalities. The brain MRI did not demonstrate evidence of striatal pathology.

**Case 5**. This participant was heterozygous for a rare *GLB1* variant (NM_000404 c.TG817-818CT, p.W273L) reported as pathogenic/likely pathogenic in ClinVar. This variant occurs in a conserved region of the gene, within a catalytic domain necessary for keratan sulfate substrate processing. Compound heterozygotes carrying p.W273L have been reported with Morquio B disease (MBD)-related skeletal dysostosis without neurological involvement; individuals typically present with predominantly skeletal manifestations. However, as MBD is an AR inherited disease, it is unclear whether one copy of this variant would be sufficient to cause parkinsonism-dystonia symptoms.

This participant was right-handed with 16 years education and parkinsonism onset between 65-70 years with tremor-predominant symptoms. There was no family history of PD. Medical conditions included hyperlipidemia, diabetes mellitus, hypertension, and mention of a “renal aneurysm” with no further information available. Neurological exam revealed rigidity, bradykinesia, and tremor and was also notable for hyporeflexia of bilateral upper extremities. In addition, the participant reported autonomic dysfunction with SCOPA-AUT score of 19 for urological and thermal regulation symptoms. MRI brain scan was unremarkable for any evidence of striatal pathologies. This presentation does not seem consistent with that reported for MBD patients carrying two copies of the *GLB1* p.W273L variant; however, it is possible that individuals with one copy would show a different phenotype.

**Case 6**. This individual was heterozygous for a rare coding change in exon 7 of the *GCDH* gene (NM_000159 c.T572C, p.M191T). This variant has been reported in individuals with glutaric acidemia type I and is classified as pathogenic/ likely pathogenic in ClinVar (accession: VCV000198396.9). However, given that variants in the *GCDH* gene show AR disease inheritance, it is unclear whether a heterozygous variant would be sufficient to cause parkinsonism-dystonia symptoms.

*GCDH* mutations cause glutaric acidemia type 1, a metabolic disease that typically manifests in infancy or early childhood with developmental delay and a progressive movement disorder that includes akinetic-rigid parkinsonism. Pathologically, it is characterized by gliosis and neuronal loss in the basal ganglia. MRI brain scans of patients with pathological mutations in the *GCDH* gene are notable for hypoplastic frontotemporal regions with widening of the sylvian fissures as well as T2 hyperintensity of the globus pallidus. However, clinical progression for this disease seems variable, with older patients with motor symptoms but normal cognition also reported in the literature.^12^

Case 6 was right-handed with 19 years education and parkinsonism onset between 45-50 years. Past medical history included osteochondrosis dissecans of left thalus and prior L4/5 disc protrusion. The case had a first-degree relative with PD. The neurological exam was notable for mild parkinsonism and bradykinesia and mild weakness of bilateral upper extremities. The MRI scan showed no abnormalities.

**Case 7**. This participant was heterozygous for a rare coding variant in exon 15 of the *ADCY5* gene (NM_001199642 c.G1816A, p.D606N). This variant is currently classified as a VUS based on the following criteria: this missense variant occurs in a gene that has a low rate of benign missense variation, and in which missense variants are a common mechanism of disease (PP2); the variant is predicted to be pathogenic (PP3), and the allele frequency is greater than expected for the disorder.

*ADCY5* variants occur in individuals initially reported as “familial dyskinesia with facial myokymia” (FDFM), but the phenotypic spectrum of this disease has been expanding and includes tremor, chorea, dystonia, and parkinsonism.^13^ While childhood onset is common, some individuals first present in adulthood. Interestingly, this disease shows an autosomal dominant pattern of inheritance, with other heterozygous missense variants segregating with disease. Other reported variants include p.A726T and p.R418W, both of which increase cAMP production in response to beta-adrenergic stimulation (gain of function).^14 15^

Case 7 was left-handed with 16 years education and parkinsonism diagnosis between 45-50 years. Past medical history was significant for iron deficiency anemia, hyperlipidemia, and anxiety. The case has a first-degree relative with PD. Main symptoms were rigidity and bradykinesia; symptoms also included postural/gait predominant symptoms and a score of 19 on the MDS-UPDRS motor scale. The neurological exam was otherwise unremarkable, including a baseline MOCA of 27. Symptom onset was significantly later than reported for other patients with *ADCY5*-related disease, though the symptom profile appears otherwise consistent.

**Case 8**. This participant was heterozygous for a rare coding variant in exon 6 of the *SGCE* gene (NM_001099400 c.G788A, p.R263H). This variant has a CADD-phred score of 28.9, meaning it is in the top 0.2% most deleterious variants in the human genome. However, it has not been reported in any dystonia patients in the literature. This variant is currently classified as a VUS by ClinVar (accession: VCV000847690.7). This missense variant has extremely low frequency in GnomAD population databases (PM2 criteria), and computational prediction tools support a deleterious effect (PP3); however, there is currently not enough evidence to support variant pathogenicity.

The *SGCE* gene has been shown to be subject to maternal imprinting, with autosomal dominant inheritance.^16 17^ Heterozygous loss-of-function variants in this gene have been identified in patients with myoclonus-dystonia syndrome. This disorder is characterized by myoclonic jerks and dystonia, though parkinsonism may be present, and many patients also have concurrent psychiatric disorders including panic attacks and obsessive-compulsive behavior.

This case was right-handed with 13 years education and parkinsonism onset between 60-65 years. The participant did not have a family history of PD. Notably, past medical history revealed dystonia and “leg cramps”. Neuropsychiatric history included the presence of an impulse-control disorder including purchasing, eating, and hobbyism. This clinical presentation is consistent with myoclonus-dystonia syndrome.

## DISCUSSION

These findings show that 13% of participants with parkinsonism and SWEDD enrolled in an observational research study of early parkinsonism harbor a predicted or reported pathogenic variant in hereditary dystonia-related genes. This supports our hypothesis that individuals with parkinsonism and SWEDD may have a unique genetic etiology different from classic PD cases, and suggests that genetic analysis should play a role in differential diagnosis for patients presenting with parkinsonism.

The occurrence of SWEDD among individuals with parkinsonism was first identified in the research setting, in clinical trials recruiting individuals with newly diagnosed PD.^18^ In 2011 the Food and Drug Administration (FDA) approved the DAT ligand for clinical use in the USA. DAT imaging was incorprated into the clinical diagnostic criteria for PD in 2015,^2^ where a normal DAT scan is specified as an exclusion criterion for the diagnosis, and DAT imaging in the clinical setting now affects diagnosis and mangement in up to a third of patients with parkinsonism.^19^ In an individual presenting with parkinsonism and with reduced dopamine tranpsorter binding on SPECT scan, the diagnosis is most likely PD or other rarer neurodegenerative parkinsonian disorders.^20^ Among those with normal DAT SPECT scans, a variety of etiologies have been posited, including vascular parkinsonism, essential tremor, or functional movement disorders.^3 20^ However, dystonic disorders mimicking PD remain high on the differential diagnosis.^21^ Dystonic disorders can lead to dystonic tremor that mimics PD rest tremor.^22^ Genetic etiologies for dystonic disorders are emerging^7 23^, and parkinsonism is part of the phenotype of many genetic dystonias. However, the contribution of pathogenic variants to the presentation of parkinsonism among those who have SWEDDs is not well studied.

We identified two participants who were heterozygous for predicted pathogenic variants in genes that have been associated with diseases with autosomal dominant inheritance patterns; one of these results has clear clinical implications. Case 8 was heterozygous for a rare coding variant in *SGCE*, p.R263H; variants in this gene have been reported in myoclonus-dystonia syndrome (DYT-SGCE; DYT11).^23^ The medical history and clinical presentation for this case align closely with reported symptoms for DYT-SGCE (DYT11), including involuntary movements and impulse-control disorder. *SGCE* p.R263H is currently classified as a VUS in ClinVar; however, given that this variant is rare in the general population, conserved, has predicted *in silico* pathogenicity, and occurs in a case with a clinically-consistent syndrome, we posit that this variant is a novel pathogenic variant for DYT11, and that DYT-SGCE (DYT11) is the etiology of the neurologic signs and symptoms in Case 8.

The second variant we identified in a gene with reported autosomal dominant inheritance was in Case 7; this individual was heterozygous for a rare coding variant in *ADCY5*, p.D606N. Other coding variants in *ADCY5* result in gain of protein function, with increased synthesis of cAMP, and have been reported in patients with dyskinesia with facial myokymia of variable severity.^14^ Molecular work is required to validate whether this variant results in gain of function. However, manifestations of parkinsonism with normal DAT scan is consistent with the diagnosis of CHOR/DYT-ADCY5, albeit a rare presentation.

We identified six additional variants with predicted pathogenicity, some of which met ACMG criteria for pathogenicity, in genes that have been associated with AR inheritance of hereditary dystonia syndromes. While the potential effects of these variants are more difficult to interpret, we posit that some of these variants may result in a milder phenotype than in patients with homozygous or compound heterozygous pathogenic variants. Another possibility is that our screen for functional coding variants may not have identified functional variants within intronic regions or adjacent (intergenic) to these genes in the affected individuals, and that some of these individuals are actually compound heterozygotes.

Strengths of this study include a large sample size of SWEDD cases and high-quality genetic data; however, this study also included several limitations. While extensive clinical and biomarker characterization was part of study activities in PPMI and was largely ascertained with standardized, validated assessments, the neurologic examination data do not specifically capture data on dystonia. In addition, medical histories and medications are collected in logs that do not capture fully discrete data. Thus, the case histories may not capture the full range of observed signs and symptoms. Additionally, the study recruited patients with recently diagnosed parkinsonism who were not expected to require symptomatic therapy for at least 6 months; cases with SWEDD were identified incidentally. This may limit the generalizability of our results. These findings require replication in the clinical setting.

These findings support that investigation of the genetic etiology of SWEDD cases is merited, as hereditary dystonia may be a significant contributing factor to misdiagnosis. Genetic screening for pathogenic variants in similar cases may uncover causal variants and aid in correct diagnosis and management.

## Supporting information

Supplemental STROBE checklist

## Data Availability

All data used in the preparation of this article were obtained from the Parkinson&#39s Progression Markers Initiative (PPMI) database (https://www.ppmi-info.org/access-data-specimens/download-data). For up-to-date information on the study, visit https://www.ppmi-info.org/.

## ACKNOWLEDGEMENTS

This work could not have been completed without the numerous talented staff supporting the PPMI study; we are very grateful for their hard work and dedication. We are also very grateful to the participants in the PPMI study, without whom this work would not be possible.

Data used in the preparation of this article were obtained from the Parkinson’s Progression Markers Initiative (PPMI) database (www.ppmi-info.org/access-data-specimens/download-data). For up-to-date information on the study, visit https://www.ppmi-info.org/.

The authors are supported by funding from the Michael J. Fox Foundation for Parkinson’s Research.

PPMI – a public-private partnership – is funded by the Michael J. Fox Foundation for Parkinson’s Research and funding partners, including 4D Pharma, AbbVie Inc., AcureX Therapeutics, Allergan, Amathus Therapeutics, Aligning Science Across Parkinson’s (ASAP), Avid Radiopharmaceuticals, Bial Biotech, Biogen, BioLegend, Bristol Myers Squibb, Calico Life Sciences LLC, Celgene Corporation, DaCapo Brainscience, Denali Therapeutics, The Edmond

J. Safra Foundation, Eli Lilly and Company, GE Healthcare, GlaxoSmithKline, Golub Capital, Handl Therapeutics, Insitro, Janssen Pharmaceuticals, Lundbeck, Merck & Co., Inc., Meso Scale Diagnostics, LLC, Neurocrine Biosciences, Pfizer Inc., Piramal Imaging, Prevail Therapeutics, F. Hoffmann-La Roche Ltd and its affiliated company Genentech Inc., Sanofi Genzyme, Servier, Takeda Pharmaceutical Company, Teva Neuroscience, Inc., UCB, Vanqua Bio, Verily Life Sciences, Voyager Therapeutics, Inc., and Yumanity Therapeutics, Inc.

## APPENDIX 1: Co-Investigators

Tanya Simuni, Caroline Tanner, Andrew Siderowf, Christopher S. Coffey, Brit Mollenhauer, Doug Galasko, Kathleen Poston, Ethan Brown, Karl Kieburtz, Roseanne D. Dobkin, Sohini Chowdhury, Todd Sherer, Mark Frasier, Kelvin Chou, Leonidas Stefanis, Stewart Factor, Maureen Leehey, Emile Moukheiber, Maria Jose Marti, Amy Amara, Stuart Isaacson, Aleksandar Videnovic, Marie Saint-Hilaire, Alberto Espay, Hubert Fernandez, Nikolaus McFarland, Arjun Tarakad, Werner Poewe, Rajesh Pahwa, Mark Lew, Yen Tai, Christine Klein, Ron Postuma, David Russel, Karen Marder, Susan Bressman, Giulietta Riboldi, Nicola Pavese, Baastian Bloem, Tiago Mestre, Michele Hu, Jean-Christophe Corvol, Nabila Dahodwala, Holly Shill, Penelope Hogarth, Ruth Schneider, Paolo Barone, Javier Ruiz-Martinez, Charles Adler, Shu-Ching Hu, David Sprecher, Robert Hauser, Nir Giladi, Connie Marras, Jan Olav Aasly, Kathrin Brockmann

