## Supplemental Methods for "Dystonia-Parkinsonism Gene Variants in Individuals with Parkinsonism and Brain Scans without Evidence for Dopaminergic Deficit (SWEDD)"

**ONLINE SUPPLEMENT**

### METHODS

#### Genetic Analyses

To identify genes associated with dystonia-parkinsonism, we referenced a list compiled by the Movement Disorder Society Task Force of 38 genes associated with isolated, combined, and complex hereditary dystonia[1]. To ensure that our results can feasibly transfer to clinical care, we also reviewed a number of commercial panels for genetic testing for parkinsonism and associated disorders, including the Fulgent Parkinson's Foundation PDGENE Panel, the Invitae Hereditary Parkinson Disease and Parkinsonism Panel, the Centogene Parkinson Disease Panel, the Prevention Genetics Parkinson Disease Panel, the Prevention Genetics Parkinson Disease and Parkinsonism Panel, the GeneDx Parkinson's Disease Panel, and the GeneDx Dystonia and Parkinsonism Panel, the GeneDx Parkinson's Disease Panel, and the GeneDx Dystonia and Parkinsonism Panel. We selected the 30 genes from the list of hereditary dystonias identified by Marras and colleagues that were present on one or more of these genetic testing panels as the final list of genes to review. This list included the following genes: *ADCY5, ATP1A3, ATP7B, CP, C19orf12, DCAF17, DDC, FA2H, FTL, GCDH, GCH1, GNAL, PANK2, PLA2G6, PRKRA, PTS, SGCE, SLC19A3, ALC30A10, SLC6A3, SPR, SUCLA2, TAF1, TH, THAP1, TIMM8A, TOR1A, TUBB4A, WDR45*, and *GLB1*.

Whole genome sequencing (WGS) VCFs for the sample were obtained from LONI (https://ida.loni.usc.edu). Exonic coding, insertions/deletions, and stop-gain variants with minor allele frequency <1% (genome aggregation database (gnomAD) genome or exome frequency in non-Finnish Europeans) or novel variants within genes of interest with at least one minor allele in SWEDD participants were identified and annotated with ANNOVAR software[2]. Genotype frequency was calculated for each variant in SWEDD, PD, and HC participants. Rare and novel variants in genes of interest that were present in SWEDD or SWEDD/PD participants but absent in controls were further assessed. Variant pathogenicity for these variants of interest was assessed, considering variant frequency in general populations, whether the DNA sequence is evolutionarily conserved, and whether the variant would be predicted to lead to abnormal gene expression or protein function. Information on these criteria were obtained from Annovar; information from a variety of variant prediction programs was included, including Sorting Intolerant From Tolerant (SIFT), Polymorphism Phenotyping v2 (PolyPhen-2), Likelihood Ratio Test (LRT), MutationTaster, FATHMM-MKL, MetaSVM, MetaLR, and Mendelian Clinically Applicable Pathogenicity (M-CAP), as well as Combined Annotation Dependent Depletion

(CADD)-phred scores (scores > 20 are considered potentially pathogenic), and Genomic Evolutionary Rate Profiling (GERP) conservation scores (scores ≥ 3 are considered conserved). Information on reported variant pathogenicity and population frequency was also obtained from ClinVar (https://www.ncbi.nlm.nih.gov/clinvar/)[3]. A final curated list of ten variants of interest was compiled based on reported pathogenicity from ClinVar, VarSome (https://varsome.com/), and Franklin (https://franklin.genoox.com/clinical-db/home) databases/programs[3 4]. The Online Mendelian Inheritance in Man (OMIM) database was referenced to obtain clinical reports from the literature for genes of interest[5].

**Clinical and Imaging Assessments**

The SWEDD cohort of PPMI was assessed at the screening/baseline visit, and one and two years afterwards. Comprehensive descriptions of the study assessments have been reported elsewhere[6 7]; briefly, they include demographics, neurological examination, medical comorbidities and medications (ascertained via review of medical condition logs and medication logs respectively, which are completed at the screening visit of the PPMI study and updated at each subsequent visit), motor signs and symptoms (Movement Disorders Society Unified Parkinson's Disease Rating Scale, MDS-UPDRS), neuropsychiatric symptoms (15-item Geriatric Depression Scale (GDS-15) and State-Trait Anxiety Inventory, STAI), cognitive testing (Montreal Cognitive Assessment, MoCA), and autonomic symptoms (SCOPA-AUT)).

-Dopamine transporter (DAT) SPECT was obtained at screening visit as previously described[18]. A binary determination of DAT binding deficit was made based on visual analyses by two expert readers.

-Structural MRI: a non-contrast enhanced, T2 weighted brain MRI using at least a 1.5 Tesla scanner and a non-contrast enhanced 3D volumetric T1-weighted brain MRI was performed at baseline for all PPMI subjects. Formal interpretation of MRIs by radiologists is not available, but MRI images, which are available at ppmi-info.org, were informally reviewed by neurologists (authors YX and LMC).
